# Entry screening and multi-layer mitigation of COVID-19 cases for a safe university reopening

**DOI:** 10.1101/2020.08.29.20184473

**Authors:** Ahmed Elbanna, George N. Wong, Zach J. Weiner, Tong Wang, Hantao Zhang, Zhiru Liu, Alexei Tkachenko, Sergei Maslov, Nigel Goldenfeld

**Affiliations:** Department of Civil Engineering, University of Illinois at Urbana-Champaign; Department of Physics, University of Illinois at Urbana-Champaign; Carl R. Woese Institute for Genomic Biology, University of Illinois at Urbana-Champaign; Computer Science, University of Illinois at Urbana-Champaign; Department of Bioengineering, University of Illinois at Urbana-Champaign; Department of Physics, Stanford University; Brookhaven National Laboratory, University of Illinois at Urbana-Champaign

**Author notes:** **Please note: this is a working document and in the interests of sharing information during a rapidly changing epidemic landscape, we are making this early version available**.

## Abstract

We have performed detailed modeling of the COVID-19 epidemic within the State of Illinois at the population level, and within the University of Illinois at Urbana-Champaign at a more detailed level of description that follows individual students as they go about their educational and social activities.

We ask the following questions:

1. How many COVID-19 cases are expected to be detected by entry screening?
2. Will this initial “bump” in cases be containable using the mitigation steps being undertaken at UIUC?

Our answers are:

1. Assuming that there are approximately 45,000 students returning to campus in the week beginning August 15, 2020, our most conservative estimate predicts that a median of 270 ± 90 (minimum-maximum range) COVID-19 positive cases will be detected by entry screening. The earliest estimate for entry screening that we report was made on July 24^th^ and predicted 198 ± 90 (68% CI) positive cases.
2. If the number of returning students is less, then our estimate just needs to be scaled proportionately.
3. This initial bump will be contained by entry screening initiated isolation and contact tracing, and once the semester is underway, by universal masking, a hybrid teaching model, twice-weekly testing, isolation, contact tracing, quarantining and the use of the Safer Illinois exposure notification app.

## Introduction

One of the many challenges arising from the COVID-19 epidemic is to develop a sustainable and safe way for educational institutions to reopen[1-9]. Here we focus on universities, rather than K-12 education, and in particular our own institution, the University of Illinois at Urbana-Champaign (UIUC). The features that make universities exciting and valuable institutions in societies world-wide --- the academic and intellectual congregation of young adults, the social opportunities, the exposure to an unprecedentedly range of activities --- are also the features that lend themselves to rapid transmission of COVID-19. Indeed, by enrolling in majors and attending a fixed set of courses, together with electives or breadth requirements, students inhabit social bubbles within a temporal small-world network whose nodes are classes and whose edges are students. Social life outside this academic network provides mixing between these social bubbles, so that one would expect transmission to occur very rapidly within a university population, perhaps more so than the population at large. In addition, the toxic combination of asymptomatic transmission, especially amongst young adults, and the inevitable bars, party and nightlife, especially in poorly ventilated settings, is highly conducive to super-spreader events. During the summer of 2020, many such instances have been reported.

In the face of these difficulties, most universities have decided to teach remotely, even though this is sub-optimal in terms of the educational experience. A few universities, including UIUC, are attempting a so-called hybrid teaching model, whereby a fraction of classes is taught online and the rest in person, but following accepted social distancing guidelines. The non-pharmaceutical interventions available to a university administration to mitigate the spread of COVID-19 include: reduced class size, universal masking, regular testing, followed by isolation, contact tracing and quarantining, and exposure notification through apps. In addition, it is critical to perform entry screening at the start of the semester to isolate students who arrive on campus already infected from their home town, many of whom will be asymptomatic or pre-symptomatic. At UIUC all of these are being deployed, in collaboration with the Champaign-Urbana Public Health District (CUPHD).

The primary question addressed in this report is: how many cases to expect to arise during entry screening? This “bump” in cases is important to anticipate in advance, because without this number as context, it is difficult for university administration to know if the cases that occur at the beginning of the semester reflect community transmission or are a temporary event that will decay once mitigation measures are deployed.

We have approached this question by estimating the bump in three different ways, each of which makes different assumptions. We discuss the merits and applicability of these different methodologies, and show that they reveal a broadly consistent picture which can be compared to universities’ reopening data. Finally, we preview, without detail, the results of agent-based models that indicate the containability of COVID-19 at UIUC through a particular set of mitigation steps. The details of these models will be published elsewhere.

## Estimate of the initial “bump”

The basic idea of the estimate is simple: we take the number of students that will be entering campus and use the prevalence of COVID-19 in the population to calculate how many will be positive. Thus:

Number of positive cases detected = (number of incoming students) × (COVID-19 prevalence)

The difficulty is that the prevalence is not directly observable so needs to be inferred somehow. We infer the prevalence in three ways: from the output of our population level models for the State of Illinois; from the daily incidence rate and the Infection Fatality Rate (IFR); and from active case data and IFR. All methods give similar results within uncertainties. We are primarily interested in the reopening of the University of Illinois at Urbana-Champaign (UIUC), but we have extended our calculations to other States and the District of Columbia and present these results below too.

For the case of UIUC, the prevalence in Champaign Country is significantly lower than that for the State as a whole, but since students return from all over the State and elsewhere, we will use the prevalence for the State not Champaign County.

### Method 1: Using computer models for the entire State

As early as July 24, 2020, we attempted to estimate the entry screening results. We have previously reported in detail on our methods to simulate the COVID-19 epidemic in Illinois and its sub-regions using a non-Markovian age-of-infection model calibrated to data provided by the Illinois Department of Public Health (IDPH) [10]. The model simulates the progression of the disease as reflected in hospital occupancy, ICU occupancy and deaths, and provides as output a prediction for the daily incidence of new infections.

**Figure 1.**
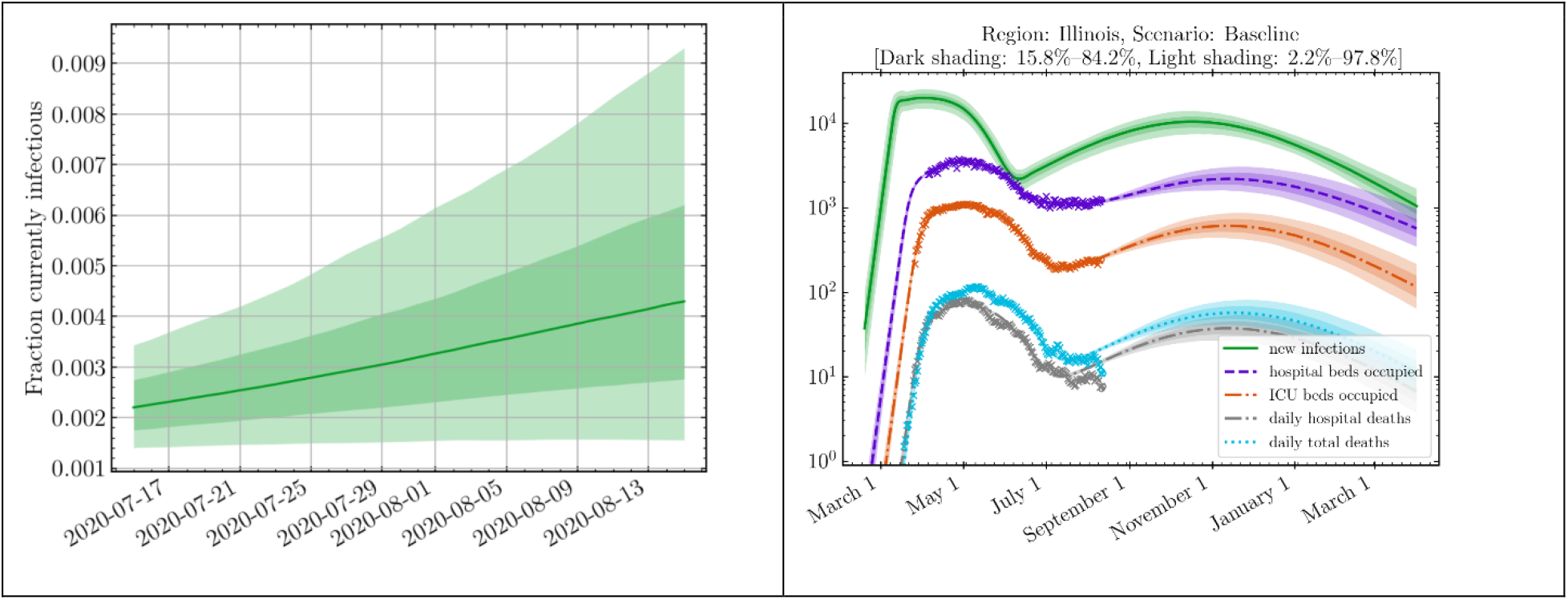
Simulation of the COVID-19 epidemic in Illinois. Left panel: projected fraction of currently infectious individuals within the State. Solid line is the median estimate, dark and light bands indicate 68% and 95% confidence intervals respectively. Right panel: projected epidemic curves for hospital occupancy, ICU occupancy, hospital deaths, all deaths and new infections (computed 8/12/2020). Crosses indicate data from IDPH.

The model predicts a prevalence of about 0.44 ± 0.2% by August 15, assuming individuals are infectious for a Gamma-distributed period with mean 5 days and standard deviation 2 days (see, e.g. [11], although such viral dynamics data are very difficult to measure precisely and were recently revised).

Thus, we estimate the number of positive cases detected = 45,000 * 0.0044 ± 0.002 = 198 ± 90

This uncertainty is for a 68% confidence interval. Using a broader estimate of uncertainty, given by the 95% confidence interval, we estimate the prevalence as lying between 0.16% and 0.92%, yielding a range of positive cases detected from 72 to 414.

### Method 2: Using daily incidence data and infection fatality ratio (IFR)

In order to refine the early estimate made three weeks in advance of the entry screening itself, we use a combination of epidemiological data and the known viral dynamics within an infectious person to provide an alternative estimate. There is no set convention for what constitutes an active case, and so we performed a different estimate of prevalence as described below. In addition, we have extended our calculation to other US States, not just Illinois.

We define prevalence *P* as:

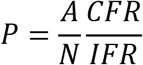

where *N* is the population of that State*, A* is active confirmed cases who will still test positive on the day of interest, *CFR* is the case fatality ratio, and *IFR* is the infection fatality ratio. We may then define the detection rate of infections *s_r_* as *s_r_* = *IFR/CFR*.

We estimate the active confirmed cases *A* who will still test positive on day *t* using the daily incidence *J_c_* as reported in [7] as follows:

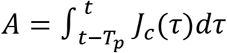

Here, we have assumed that people who have tested positive *T_p_* days ago will still test positive on the current day *t*. The above integral may be further simplified using the following approximation

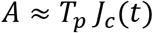

Here *J_c_*(*t*)is the confirmed daily incidence on day *t* computed using a 7-day moving average. The above approximation is likely to be an overestimate in states with growing epidemic while being an underestimate in states with shrinking epidemic. However, the error in either cases is acceptable within limits of uncertainty of different parameters characterizing the epidemic.

To estimate the Case Fatality Ratio, we use the following formula:

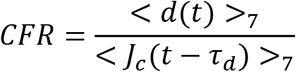

Here <*d*(*t*)>_7_ is the 7-day average of daily death at time *t* and <*J_c_*(*t*−*τ_d_*)>_7_ is the 7-day average of confirmed daily incidence from *τ_d_* days in the past. This shift is important to account for delays between onset of symptoms (which generally has been found to be close to the day of testing) and death. We take *τ_d_* = 21 days.

For the state of Illinois we have the following estimates. Several studies [2,5,6] suggest that people may test positive for up to *T_p_* = 21 days after the onset of symptoms. The confirmed 7-day average of the daily incidence in the State on August 21,2020 is 1903. Thus the active confirmed cases who may test positive on that day is approximately given by 21 × 1903 = 39963.

The case fatality ratio CFR on August 21,2020 is estimated as follows. <*J_c_*(*t*−21)>_7_=1488, <*d*(*t*)>_7_=19.1. Accordingly, CFR = 19.1/1488 = 1.28%.

Assuming an IFR = 1.0% we thus estimate that *P* = 0.4%.

Assuming an IFR = 0.5% we estimate that *P* = 0.8%.

The above estimates for prevalence in the State range between 0.4% and 0.8%. This range is consistent with the estimates from our age of infection model (Method 1) that give a 95% credible interval for the range of prevalence between 0.16% and 0.94%.

Using the above methodology, we present estimates for prevalence in the 50 States and the District of Columbia in the Supplementary Information. Figure 2 shows the range of these values. The red dash in each box corresponds to the median value. For the state of Illinois, the median predicted value for the prevalence is 0.6%.

**Figure 2.**
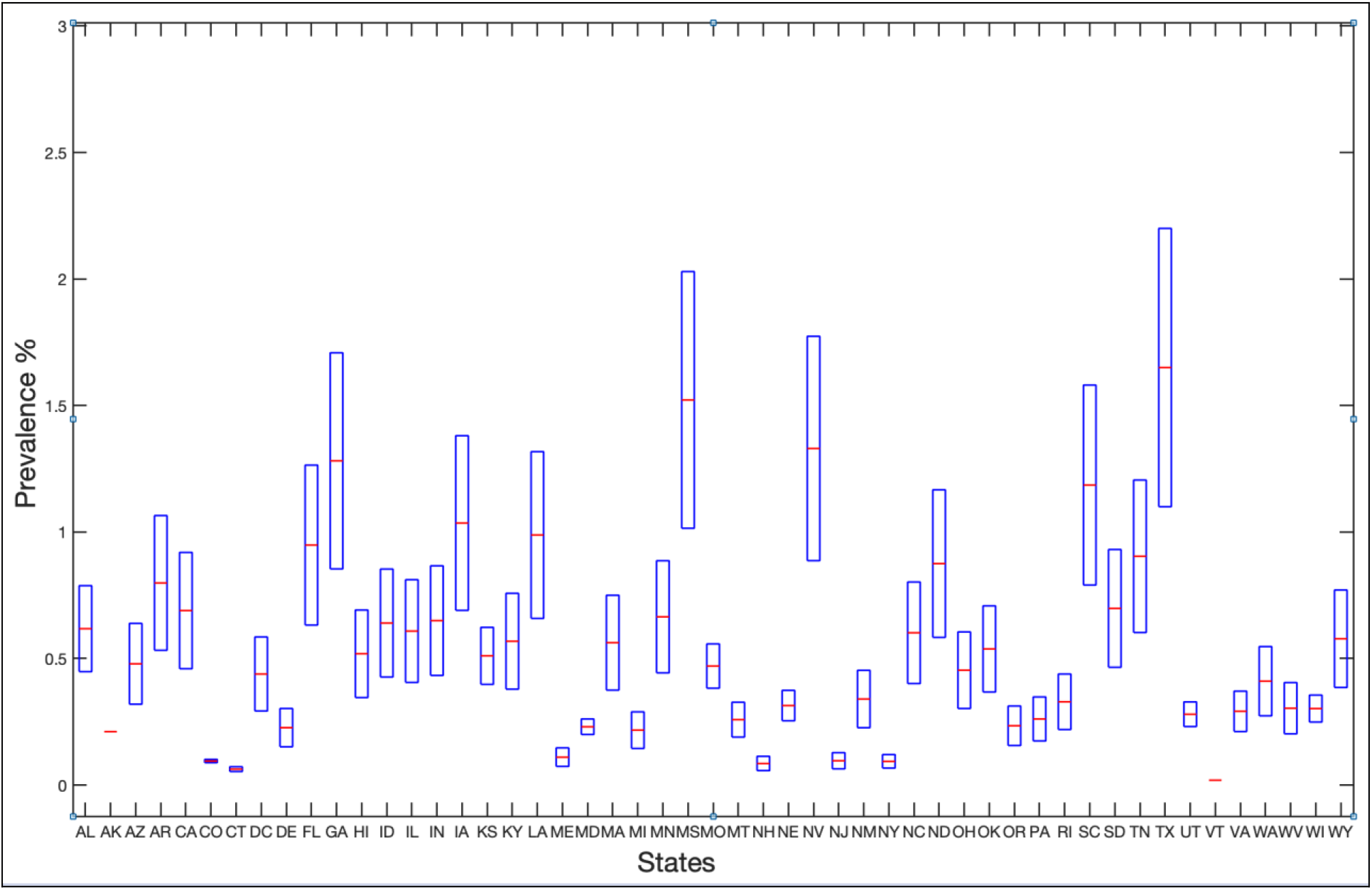
Estimated prevalence in the 50 states and the District of Columbia computed using Method 2 on August 21, 2020. Details are provided in the Supplementary Information. The median value is indicated by the red dash, the box delineates the maximum and minimum values.

### Method 3: Using active cases data and infection fatality ratio (IFR)

In order to give an independent estimate, we can use epidemiological data available from the IDPH website (https://www.dph.illinois.gov/covid19). The idea is to work backwards from the known COVID-19 deaths to infer the total number of infections. This number would be the same as the number of COVID-19 cases measured in the State, if the State was able to test everyone all the time. In practice, this is impossible, so by comparing the total number of infections, we can estimate the detection rate. Once we have the detection rate, we can work out the actual number of infected people in Illinois right now from the number that have actually been detected and confirmed COVID-19 positive. Dividing by the population of the State gives the prevalence.

On Aug 14, 2020, the total number of COVID-19 deaths was 7,721. The IFR is usually taken to be 0.7% and is almost certainly within the range 0.5%-1.0% according to the World Health Organization [3]. This means that the total number of people who have been infected in the State of Illinois is 7721/0.007 = 1,103,000. IDPH has detected 202,691 confirmed cases during the course of the epidemic, so the average detection rate is 202,691/1,103,000 = 18.3%. This is a crude estimate because the testing has increased from < 5000 per day at the early stages to nearly 50,000 tests per day, and the severity of the epidemic has grown, declined and started to grow again in a second wave. A more reliable estimate for the detection rate may be computed using the case fatality ratio CFR on Aug 14, 2020 which is estimated to be equal to 1.2% (See Table in Supplementary Information). Accordingly, the detection rate is 0.7/1.2 = 58%. For these cases, the recovery rate is 95% according to IDPH [4], so the number of active confirmed cases known right now in Illinois might seem to be 0.05 × 202,691 = 10,134. However, this is misleading, because the recovery rate is defined (in Illinois only) as those cases whose specimen collection occurred more than 42 days ago, and who have not died, divided by the sum of such cases and the number of COVID deaths. The implications of this definition are the following. The number of active confirmed cases is actually, according to the IDPH definition, 202,691 – 20 × 7721 = 48,271. The time period of 42 days used in this definition is not supported from the perspective of the viral dynamics. In fact, in prior work on temporal viral dynamics [12], the virus was not detected beyond 33 days after symptom onset. Furthermore, according to these studies, the virus is likely to be undetected in most samples beyond 21 days. Thus only a fraction of these 48,271 cases are likely to test positive again on Aug 14, 2020. We estimate this fraction, guided by the current state of the art of viral dynamics, to be in the range of 21/42 x 48,271 = 24,135 to 33/42 x 48,271 = 37,927. These estimates ignore the cases that have not been detected. To work out the actual number of people who may test positive on Aug 14, 2020, we correct by dividing these number by the detection rate 58% to get a range between 41,612 and 65,391. Finally, we estimate the prevalence by dividing by the population of the State to get 0.33% < *P <* 0.51%. Again, this range is consistent with the ones estimated in Methods 2 and 3.

We summarize the estimates for the number of entry-screened cases in the table below.

**Table 1:** Summary of estimates for N_ES_, the number of cases detected by entry screening at the University of Illinois at Urbana-Champaign using three different methods of estimation as described in the text.

| Method | Result |  | Nature of uncertainty interval |
| --- | --- | --- | --- |
| | $N_{ES}$ | $P \%$ | |
| Method 1 | $N_{ES} (50\%) = 198$<br>$72 < N_{ES} < 414$ (95% CI)<br>$108 < N_{ES} < 288$ (68% CI) | $P (50\%) = 0.44$<br>$0.16 < P < 0.92$<br>$0.22 < P < 0.66$ | Projections made on July 24<sup>th</sup>, 2020 for prevalence on Aug 12<sup>th</sup>, 2020 using the age of infection model. |
| Method 2 | $N_{ES} (50\%) = 270$<br>$180 < N_{ES} < 360$ (Min-Max) | $P (50\%) = 0.6$<br>$0.4 < P < 0.8$ | Uncertainty in Infection Fatality Ratio |
| Method 3 | $N_{ES} (50\%) = 189$<br>$149 < N_{ES} < 230$ (Min-Max) | $P (50\%) = 0.42$<br>$0.33 < P < 0.51$ | Uncertainty in the time duration over which virus may continue to be detectable post symptom onset |

## Effectiveness of mitigation

We have simulated the spread of COVID-19 on campus using agent-based modeling, following each student as they attend university activities and participate in off-campus socializing. Full details will be given elsewhere. For now we simply note some of the assumptions of our models, some of which are more conservative than what is actually the case: in other words, we are likely over-estimating the severity of the epidemic and under-estimating some of the mitigation measures. At UIUC, these measures are known collectively as “SHIELD”, and implemented in partnership with the Champaign-Urbana Public Health District (CUPHD).

- SARS-CoV-2 is assumed to be transmitted through local interactions and aerosols [13-19]
- Universal masking within University buildings. However, we do not assume that students will be wearing masks in restaurants or bars or other off-campus premises. We are thus underestimating the effectiveness of masks.
- We follow all the students who had classes in Fall 2019 as a representation of the University activity: this is about 45,000 students. The actual number may be much less in Fall 2020.
- All classes with more than 50 students are online. This represents about 40% of the total number of classes. We assume the rest are in person. In fact, UIUC policy allows professors to decide how they wish to teach, and with the publicly stated figure of 1/3 in-person classes, we are under-estimating the number of online classes and over-estimating the infection spread within the University.
- We assume 2 tests per week, using UIUC’s saliva testing protocol [20].
- We assume contact tracing (CT) works backwards to trace all interactions between the index case and contacts that are longer than 15 minutes, closer than 6 feet, within the 48 hours previous to testing. Contact tracing effectiveness is assumed to be 50%: half the contacts are missed.
- We assume exposure notification (EN) uses a privacy-preserving protocol that only provides an alert for a nearby contact detected via Bluetooth if the duration was longer than 2 hours. This is to ameliorate the problem of false-positives and weights the detected events only to those where there is a high likelihood of exposure (so-called “risk-weighted” exposure notification). We compare our own algorithm for risk-weighted exposure notification (EN), which will be reported elsewhere, to other existing exposure notification applications that are currently used worldwide. These applications include (i) the German Corona Warn App (CWA), which generates exposure notifications with the risk of exposure weighted by the idealized infectiousness profile of a typical Covid-19 patient as reported in [11]; and (ii) the Google/Apple Decentralized Privacy Preserving Proximity Tracing (DP3T) application which generates exposure notification for any interaction longer than 15 minutes with an individual who has tested positive.
- Students move on a schedule between a variety of zones: home, lectures, library, coffee shops, restaurants, bars/parties on a schedule and infect each other with epidemiological characteristics derived from the literature, especially [11,21,22]. Masks are assumed only in University buildings, not in social life --- a worst case scenario.

In Figure 3 we show the results of our agent-based model for the start of the semester, assuming that 200 agents arrive on campus as test positive. The curve labelled “infected” shows the active cases, and the curve labelled “isolated” shows the number of cases detected by our frequent testing protocol and isolated. The isolated cases is not congruent to the active cases for two reasons: first there is a delay due to the testing protocol, and second because of the false negative rate assumed in the testing of 11%. The initial “bump” is clearly visible but decays over about 20 days. The speed with which this bump decays is sensitive to the delays in getting testing results returned. In the figure, we used a 5 hour delay, which is what we have anticipated for the Illinois pipeline.

**Figure 3.**
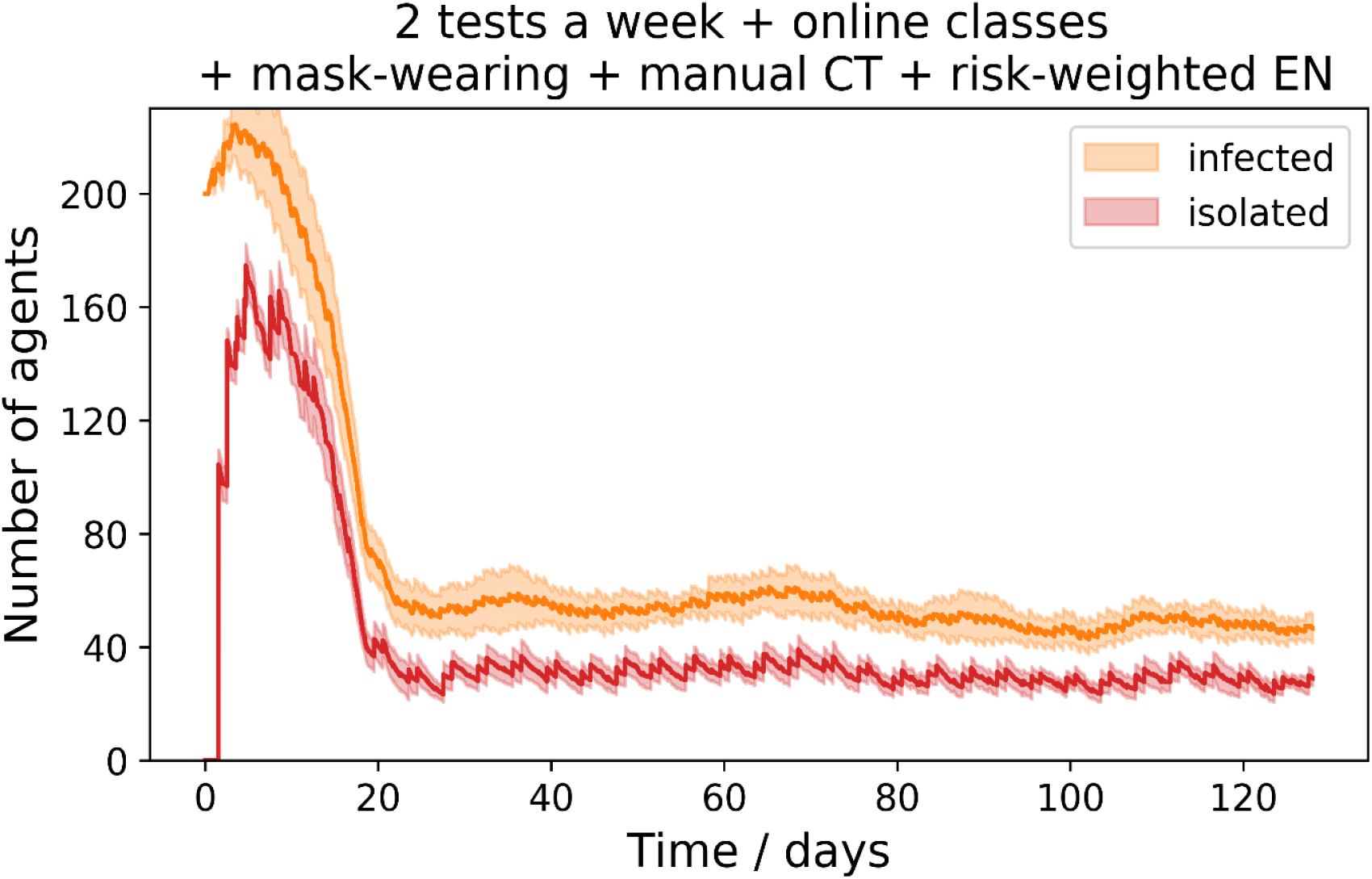
Simulation of 45,000 student epidemic curves during the semester, starting from an initial infection of 200 students. The figure shows the impact of multiple mitigation strategies in containing the epidemic at a steady state, and its effectiveness in bringing under control the initial “bump” in cases. Simulations are repeated 10 times and median and standard deviation is shown.

In Figure 4, we show additionally the number of new cases expected and the number of active cases.

**Figure 4.**
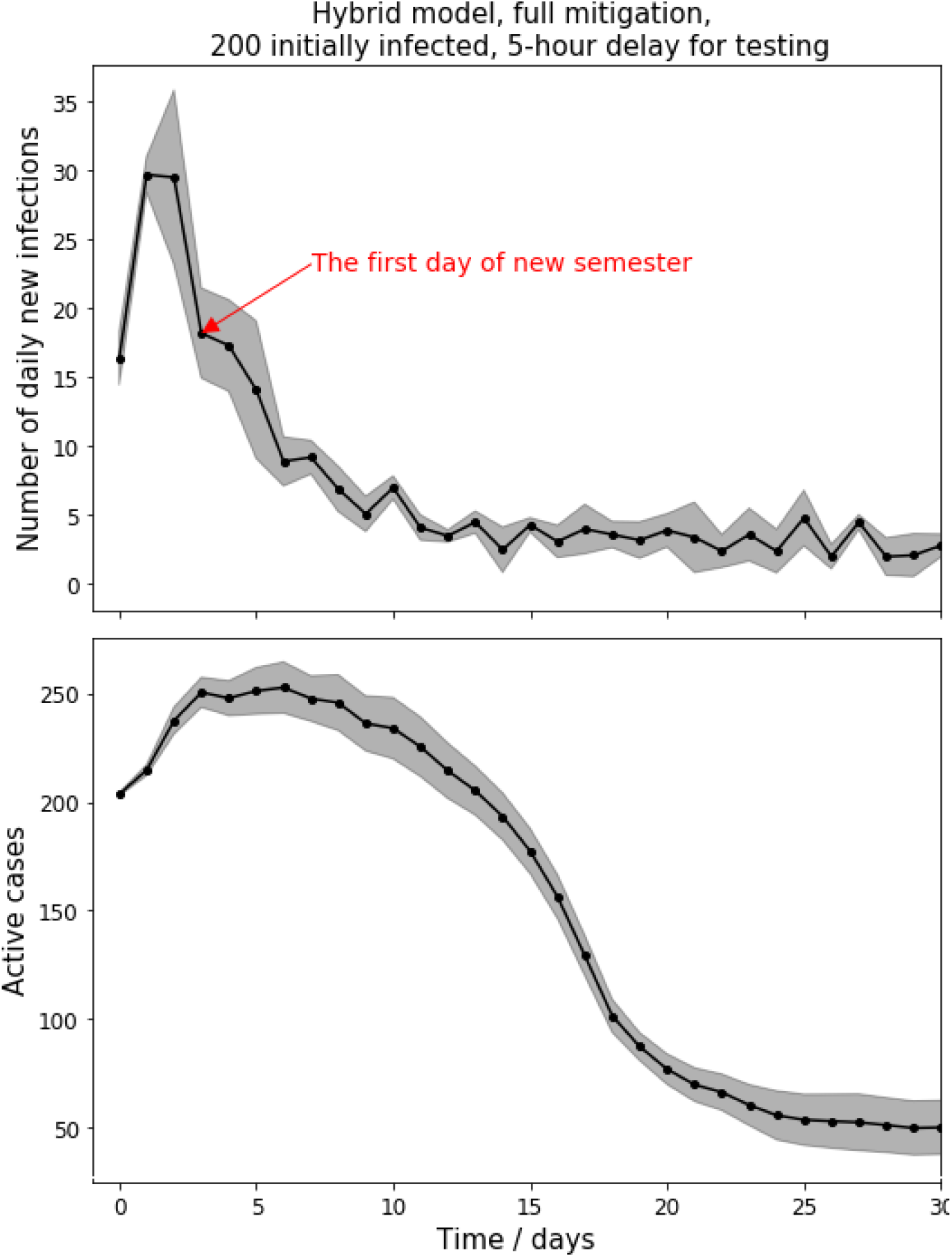
Simulation of 45,000 student epidemic curves during the semester, starting from an initial infection of 200 students. Top: Daily number of new cases. Bottom: Active cases. Simulations are repeated 10 times and median and standard deviation is shown.

Over the semester the total number of infected agents is shown in Figure 5, starting with unchecked disease progression, and layering on successive mitigation modalities. We only show the effect of testing, masks, online classes and manual contact tracing. Exposure notifications can in principle reduce the number of infections but not by a number that is statistically significant if the other modalities are working. This would be different, for example, if manual contact tracing was inoperative: exposure notification on its own is about as effective as manual contact tracing (but only if there is > 60% adoption of the exposure notification app).

**Figure 5.**
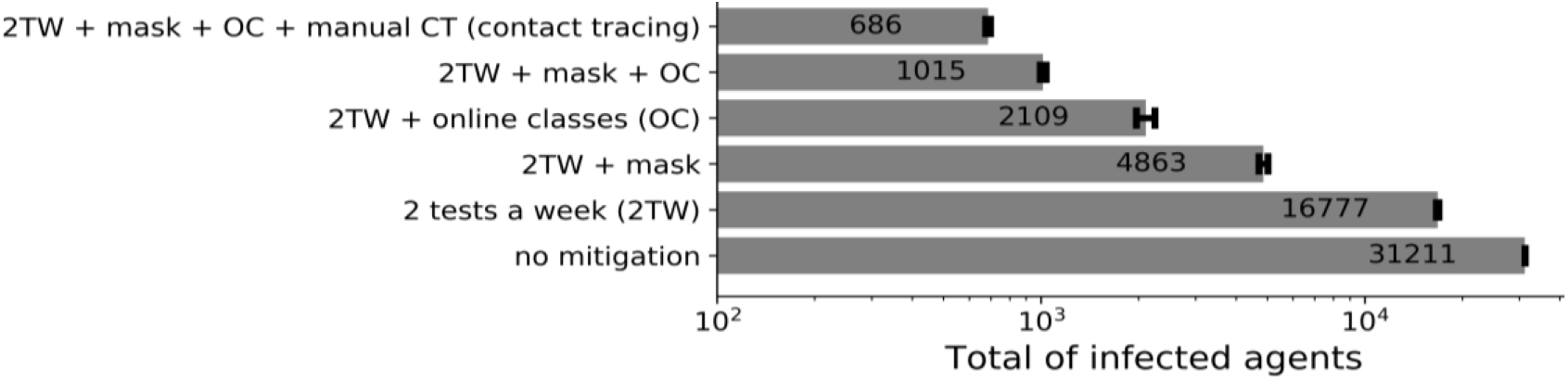
Total number of infected students during the semester under a variety of mitigation “bundles”, including different protocols for exposure notification. This number includes the initial 200 infected students. The results shown here are for a particular transmission model, and are only weakly sensitive to model assumptions, such as aerosol transmission and other disease characteristics.

In Figure 6, we show the maximum number of infected cases who are isolated at any one time and the number of students quarantined as a result of contact tracing. The maximum isolation size is dominated by the early period of the semester when there are 200 imports to campus (this number is not exactly equal to 200, because of the false negative rate of the UIUC saliva test). In steady state, after the initial burst, the number is about 50.

**Figure 6.**
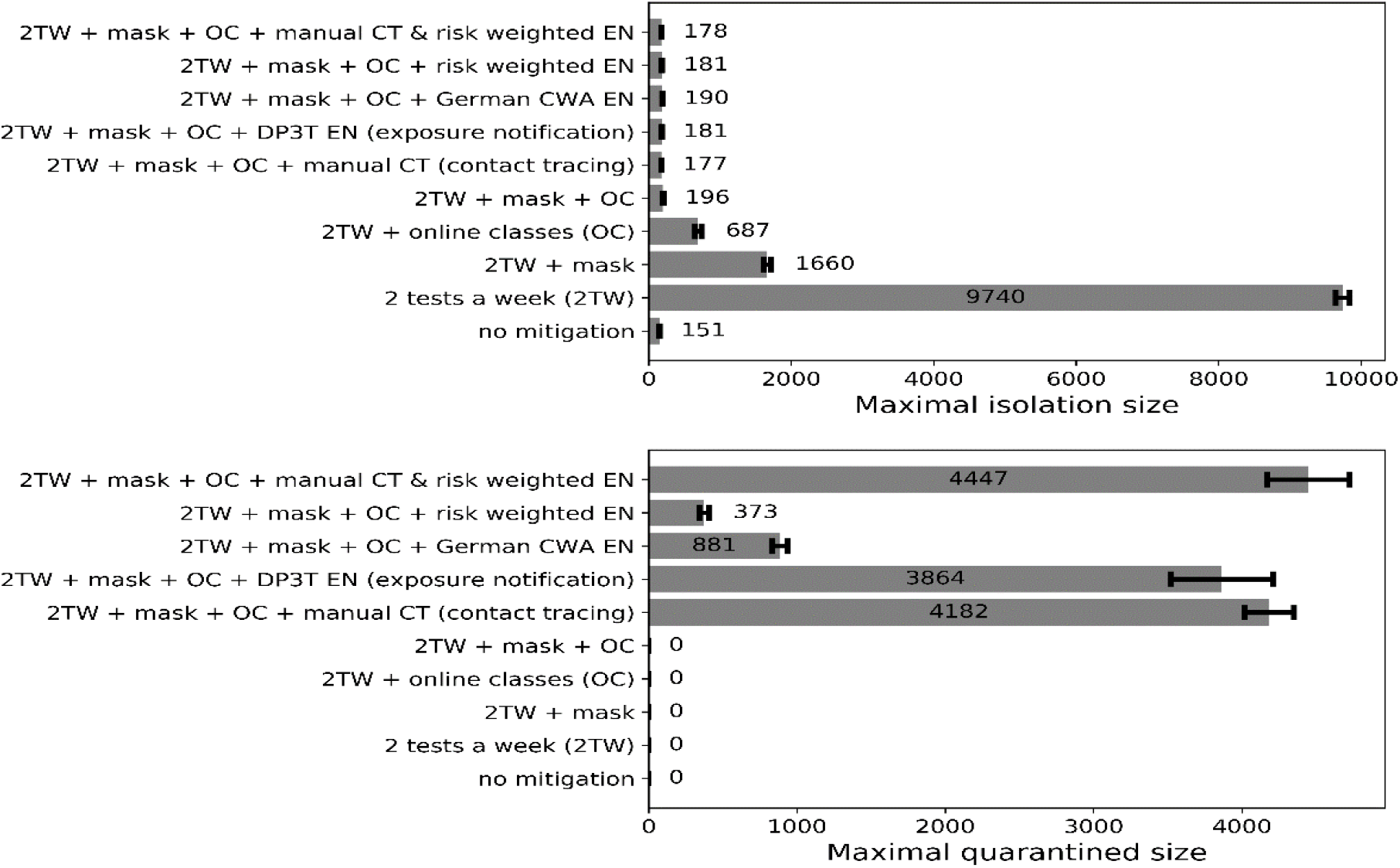
Maximum number of isolated COVID-19 confirmed cases and quarantined contacts, under a variety of mitigation measures.

## Conclusion

When students return to campus, a University must be prepared for there to be a significant number of detected COVID-19 confirmed cases. These individuals must be isolated, and asked to participate in contact tracing, in accord with public health district policies. It is essential that these be followed and especially that returning students work closely with public health officials to break the chains of transmission. Non-compliance in this can be very destructive potentially or prolong the initial burst.

Overall, multi-layered mitigation including regular testing of the entire university population, universal masking, a hybrid teaching model, isolation, contact tracing and quarantine, and exposure notification can be very effective in enhancing student safety. By extension, a contained epidemic within the student population combined with a low prevalent state in the community will help provide as safe an environment as possible for all.

## Data Availability

N/A

## Acknowledgments

We gratefully acknowledge discussions with Mark Johnson at Carle Hospital and Awais Vaid at Champaign-Urbana Public Health District. We gratefully acknowledge helpful discussions with members of the UIUC SHIELD team, especially Marty Burke, Paul Hergenrother, Sanjay Patel, Rebecca Lee Smith, William Sullivan, Nicholas Vance. Our calculations would have been impossible without the data kindly provided by the Illinois Department of Public Health through a Data Use Agreement with Civis Analytics. This work was supported by the University of Illinois System Office, the Office of the Vice-Chancellor for Research and Innovation, the Grainger College of Engineering, and the Department of Physics at the University of Illinois at Urbana-Champaign. Z.J.W. is supported in part by the United States Department of Energy Computational Science Graduate Fellowship, provided under Award No. DE-FG02-97ER25308. This work made use of the Illinois Campus Cluster, a computing resource that is operated by the Illinois Campus Cluster Program (ICCP) in conjunction with the National Center for Supercomputing Applications (NCSA) and which is supported by funds from the University of Illinois at Urbana-Champaign. This research was partially done at, and used resources of the Center for Functional Nanomaterials, which is a U.S. DOE Office of Science Facility, at Brookhaven National Laboratory under Contract No.~DE-SC0012704.

